# Salivary IgG antibody response to SARS-CoV-2 as a non-invasive assessment of immune response. Differences between vaccinated children and adults

**DOI:** 10.1101/2024.10.18.24315700

**Authors:** María Noel Badano, Irene Keitelman, Matías Javier Pereson, Natalia Aloisi, Florencia Sabbione, Patricia Baré

## Abstract

**Background:** Studies comparing systemic and salivary antibody responses against SARS-CoV-2 between children and adults show conflicting results. Furthermore, it is still unclear whether salivary antibody testing could be a non-invasive approach to evaluate the humoral immune response.

**Methods:** anti-SARS-CoV-2 IgG antibodies were measured in blood and saliva sample pairs from vaccinated adults to investigate whether salivary antibody response could be a non-invasive assessment of immune response. Salivary antibody levels were also compared between vaccinated children and adults to investigate local antibody responses.

**Results:** Salivary IgG antibody response against SARS-CoV-2 largely reflects the systemic response in vaccinated adults. Salivary and systemic antibody concentrations were higher in vaccinated adults who had been infected, received schedules including mRNA-based vaccines, had more exposures, and a shorter time from last exposure. Salivary antibody detection was associated with schedules including mRNA-based vaccines, time from last exposure, and systemic antibody concentrations. Vaccinated children showed higher salivary antibody concentrations than adults. This difference remained when comparing antibody levels between children and adults under equal conditions (vaccination schedules, number of exposures, time from last exposure, COVID-19 history). Younger age, number of exposures, schedules including mRNA-based vaccines, and shorter time from last exposure were associated with salivary antibody levels in a multivariable linear regression analysis (*p*< 0.0001).

**Conclusions:** Salivary antibody determination against SARS-CoV-2 could be a non-invasive assessment of the short-term immune response in adults with multiple exposures. Furthermore, the stronger salivary antibody response in children suggests that local immune protection may differ between children and adults, contributing to different outcomes.

## Introduction

While several advances have been made in the understanding of SARS-CoV-2 infection and its associated pathology, certain aspects related to the immune response remain uncertain. Furthermore, the lower availability of blood samples from children coupled with the late administration of COVID-19 vaccines in this population led to less knowledge about their immune response following infection and vaccination in comparison to adults.

It is still debatable whether children and adults show similar systemic humoral immune responses against SARS-CoV-2 after infection and vaccination. Studies have reported similar [1, 2], lower [3, 4] or stronger and longer-lasting [5, 6] systemic antibody responses to SARS-CoV-2 infection in children than adults. There are also conflicting results regarding antibody responses following vaccination, with studies showing higher [6, 7], similar [8–10] or lower [4] SARS-CoV-2-specific antibody levels in children compared to adults.

Information about the mucosal immune response against SARS-CoV-2 in children is limited. Using saliva-based tests, the prevalence of salivary anti-SARS-CoV-2 IgG antibodies in unvaccinated and vaccinated children has been studied [11–13], further showing that salivary antibody levels increase following infection or household contact exposure [12, 14] and vaccination [13]. However, studies comparing salivary antibody responses against SARS-CoV-2 between children and adults have yielded contradictory results, showing higher [15] or lower [4] salivary antibody levels in children than in adults.

On the other hand, results from studies comparing systemic and salivary humoral immune responses to SARS-CoV-2 are inconsistent. While several studies have shown a positive correlation between systemic and salivary anti-SARS-CoV-2 IgG antibody levels in both adults [16–18] and children [11, 15], other studies have shown a weak correlation [19, 20]. We have also previously shown that SARS-CoV-2 breakthrough infections in vaccinated adults boost antibody levels not only in the blood but also in the salivary compartment [21]. Therefore, although several studies have demonstrate the validity of assessing specific salivary antibodies as indicators of seroconversion after natural infection or vaccination [16–18, 22–23], it remains unclear whether saliva could be suitable as a non-invasive alternative to blood for monitoring antibodies against SARS-CoV-2.

In this work we investigated and compared the systemic and salivary IgG antibody responses against SARS-CoV-2 in vaccinated adults, to analyze whether the determination of salivary antibodies against SARS-CoV-2 could be a non-invasive approach to evaluate the humoral immune response following infection or vaccination. Salivary antibody levels were also compared between vaccinated children and adults to analyze whether the specific humoral immune response differed within the salivary compartment.

## Materials and methods

### Study Design, Participants and Samples

Since the beginning of the COVID-19 pandemic, we conduct an observational prospective cohort study to investigate the humoral immune response against SARS-CoV-2 after infection and vaccination. Adult participants were healthcare workers from our institution, the Academia Nacional de Medicina. Child participants were sons and daughters of healthcare workers and their schoolmates. SARS-CoV-2 diagnoses were conducted in adults and children with COVID-19 symptoms and in some household members without symptoms. If a household member tested positive for SARS-CoV-2 and no isolation measures were taken, all other household members were considered exposed (household contacts), except those who were subsequently diagnosed. SARS-CoV-2 infection was confirmed by RT-PCR in nasopharyngeal/oropharyngeal swabs or in saliva samples. Children and adults were monitored and information regarding demographic characteristic (date of birth, sex), vaccination (dates and schedules), COVID-19 history (date and presence of symptoms in positive cases and household contacts) was completed in an online questionnaire by adults and parents.

In this study, blood and saliva sample pairs (*n* = 131) from adults who had received two or three vaccine doses collected between December 2021-July 2022 were included. Saliva samples (*n* = 88) from children up to 18 years old who had received two or three vaccine doses collected between February-August 2022 were also included. The samples included were obtained from uninfected adults and children, as well as from those who became infected or were household contacts at any stage of vaccination. All subjects with confirmed past SARS-CoV-2 infection had mild disease based on the World Health Organization classification [24]. Information regarding demographic characteristic, COVID-19 history, vaccination schedules, number of exposures (through vaccination, infection or household contact exposure) and time between last exposure and sample collection is detailed in Table 1. Vaccines from the same platform were administered on similar dates and with similar interval time between doses. This study was approved by the Academia Nacional de Medicina Ethics Committee. Written informed consent was obtained from adult participants and from parents for children participants.

**Table 1.** Characteristics of children and adults.

|  | **Children**<br>**(n=88)** | **Adults**<br>**(n=131)** |
| --- | --- | --- |
| **General characteristics** |  |  |
| Age, median (range), y | 10 (4–17) | 45 (27-83) |
| Sex |  |  |
| Female, No. (%) | 40/88 (45) | 91/131 (69) |
| Male, No. (%) | 48/88 (55) | 40/131 (31) |
| **COVID-19 history** |  |  |
| Uninfected, No. (%) | 35/88 (40) | 67/131 (51) |
| Confirmed past SARS-CoV-2 infection, No. (%) | 16/88 (18) | 56/131 (43) |
| Household contacts, No. (%) | 37/88 (42) | 8/131 (6) |
| Infected/household contacts with COVID-19 compatible symptoms, No. (%) | 32/53 (60) | 55/64 (86) |
| Infected/household contacts exposed to pre-Omicron variants | 15/53 (28) | 17/64 (27) |
| Infected/household contacts exposed to Omicron variant | 38/53 (72) | 47/64 (73) |
| **Vaccination schedules non-mRNA-based** |  |  |
| BBIBP-CorV x 2, No. (%) | 46/88 (52) | 5/131(4) |
| ChAdOx1 nCoV-19 x 2, No. (%) |  | 5/131(4) |
| Sputnik V x 2, No. (%) |  | 1/131(1) |
| BBIBP-CorV x 2 + ChAdOx1 nCoV-19 x 1, No. (%) | – | 66/131 (50) |
| Sputnik V x 2 + ChAdOx1 nCoV-19 x 1, No. (%) | – | 10/131 (8) |
| ChAdOx1 nCoV-19 x 3, No. (%) | – | 7/131 (5) |
| **Vaccination schedules mRNA-based** |  |  |
| BNT162b2 mRNA x 2, No. (%) | 7/88 (8) | – |
| mRNA-1273 x 2, No. (%) | 1/88 (1) | – |
| BNT162b2 mRNA x 3, No. (%) | 17/88 (19) | – |
| BNT162b2 mRNA x 2 + mRNA-1273 x 1, No. (%) | 4/88 (5) | – |
| BBIBP-CorV x 2 + BNT162b2 mRNA x1, No. (%) | 11/88 (13) | 8/131 (6) |
| BBIBP-CorV x 2 + mRNA-1273 x 1, No. (%) | 2/88 (2) | 4/131 (3) |
| Sputnik V x 2 + BNT162b2 mRNA x1, No. (%) | – | 8/131 (6) |
| ChAdOx1 nCoV-19 x 2+ BNT162b2 mRNA x1, No. (%) | – | 4/131 (3) |
| Sputnik V x 1 + mRNA-1273 x 2, No. (%) | – | 13/131 (10) |
| **Number of antigen exposures** |  |  |
| Two vaccine doses (2X), No. (%) | 23/88 (26) | 8/131 (6) |
| Two vaccine doses plus one infection/household contact exposure (3X), No. (%) | 27/88 (31) | 3/131 (2) |
| Three vaccine doses (3X), No. (%) | 12/88 (14) | 59/131 (45) |
| Two vaccine doses plus two household contact exposures (4X), No. (%) | 4/88 (4) | – |
| Three vaccine doses plus one infection/household contact exposures (4X), No. (%) | 22/88 (25) | 61/131 (47) |
| Time from last antigen exposure and sample collection, median (range), days | 76 (21–270) | 50 (21–234) |
| Time from last antigen exposure and sample collection, median (range), days | 58 (21–270) |  |

BBIBP-CorV (Sinopharm); ChAdOx1 nCoV-19 (University of Oxford/AstraZeneca); BNT162b2 mRNA (Pfizer–BioNTech); mRNA-1273 (Moderna).

### Sample collection and processing

Plasma or serum samples were obtained immediately after centrifugation of the peripheral blood and stored in aliquots at −20°C until use. For saliva sampling, individuals provided their first saliva of the day by spitting into a tube, ensuring they had not consumed any food or drink, brushed their teeth, or engaged in other activities prior to collection that could introduce variability. Saliva samples were centrifuged at 17,000 x *g* for 10 min (4°C) and the supernatant was stored at −20°C until use.

### SARS-CoV-2 Antibody ELISA

Detection of anti-spike SARS-CoV-2 IgG antibodies in both blood and saliva was performed by ELISA (COVIDAR-IgG, Laboratorios Lemos S.R.L, Buenos Aires, Argentina). The assay plates are coated with a purified mixture of the spike protein and the receptor binding domain (RBD) of SARS-CoV-2 from the original Wuhan viral variant (GenBank: MN908947). Determination of specific antibodies in blood was performed following the manufacturer’s instructions [21].

Assessment of salivary antibodies was performed with the conditions we had previously established [21]. Basically, salivary IgG anti-spike antibodies were determined as plasma ones, without performing the first sample dilution. As pre-pandemic samples were unavailable, negative controls comprised PCR-negative saliva samples from early stages of the pandemic obtained before vaccination. Antibody concentrations in binding antibody units (BAU) per mL (BAU/mL) were obtained by interpolating the OD 450 nm values of the samples into a calibration curve performed with the provided standard (400 BAU/mL). Each sample was properly diluted to fit an OD 450 nm within the linear range of the calibration curve.

### Statistical Analysis

Data analyses were performed using the GraphPad 10.2.3 Prism software (GraphPad Software, San Diego, CA, USA). Geometric mean concentrations (GMC) of specific antibody levels with 95% confidence intervals (95% CI) were calculated. Antibody levels were analyzed according to variables previously reported to be associated with systemic anti-SARS-CoV-2 IgG antibody levels (age, sex, number of exposures, vaccination schedules, SARS-CoV-2 infection and time between most recent antigen exposure and sample collection). Antibody levels between two groups were compared with Mann–Whitney test. A multivariate linear regression model was performed including these variables as independent variables and salivary antibody levels as the dependent variable. The linear regression coefficients (β) with 95% CI were calculated. The Spearman coefficient of rank correlation was used to assess the correlation between specific salivary and blood antibodies. In all cases, a value of *p*< 0.05 was considered indicative of a significant difference.

## Results

### IgG specific salivary and blood antibody responses in vaccinated adults

Specific antibodies were detected in 100% of blood samples (GMC BAU/mL, 95% CI: 3589, 2800−4601) and in 84% of saliva samples (GMC BAU/mL, 95% CI: 78.4, 53.9−114.0). A positive correlation was observed between levels of specific antibodies in blood and saliva (*r*= 0.7, *p*<0.0001) (Figure 1A). Subjects with higher systemic antibody levels (≥GMC: 3589 BAU/mL) also showed higher salivary antibody concentrations compared to those with lower systemic antibody levels (GMC BAU/mL, 95% CI, Saliva: 300.3, 227.1−397.2 *vs* 24.3, 12.3−48.1; *p*< 0.0001) (Figure 1B). Furthermore, higher systemic antibody levels were found in subjects with detectable salivary antibodies compared to those without detectable salivary antibodies (GMC BAU/mL, 95% CI, Blood: 4529, 3556−5767 *vs* 964.8, 515.5−1806; *p*< 0.0001) (Figure 1C).

**Figure 1.**
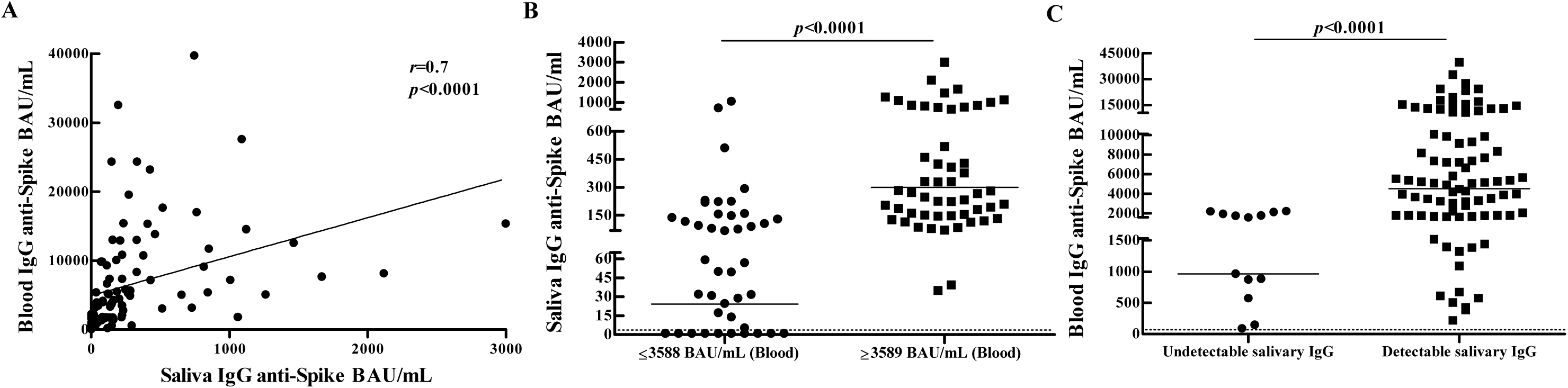
Systemic and salivary antibody responses against SARS-CoV-2 in vaccinated adults. (A) Correlation between systemic and salivary SARS-CoV-2-specific IgG responses. Antibody levels were measured in blood and saliva sample pairs from vaccinated adults adults (*n* = 131). Correlation between specific salivary and blood antibodies was analyzed by the Spearman rank correlation test. Spearman correlation coefficient (*r*) and *p*-value are indicated. (B) Salivary anti-SARS-CoV-2 antibody concentrations were compared between subjects with higher systemic antibody levels (≥GMC: 3589 BAU/mL) and subjects with lower systemic antibody levels (≥GMC: 3588 BAU/mL). (C) Comparison of systemic anti-SARS-CoV-2 antibody levels between subjects with or without detectable salivary antibodies. IgG anti-spike antibody concentrations (BAU/mL) with geometric means are shown. Dotted line indicates the assay detection limit (4.03 BAU/mL). *p* values were determined by Mann-Whitney test. Samples were assayed in duplicate and the results shown are representative of one of two independent experiments performed.

Therefore, we analyzed how variables previously reported to be associated with systemic anti-SARS-CoV-2 IgG antibody levels (see Materials and methods for details), relate to antibody levels in both the blood and the salivary compartment. Similar results were observed in the blood and in the salivary compartment. The results of the variables where significant differences in antibody levels were observed are shown. As similar results were observed among subjects who received one or two doses of mRNA-based vaccines, they were combined into a single group (non-mRNA + mRNA (3d)) for results presentation. Levels of anti-spike SARS-CoV-2 IgG antibodies were higher in subjects who received three doses of schedules that included mRNA-based vaccines (non-mRNA + mRNA (3d)) compared to those in individuals who received three doses of schemes not including mRNA-based vaccines (non-mRNA (3d)) (GMC BAU/mL, 95% CI, Blood: 8745, 6102−12533 *vs* 2574, 1949−3398; *p*< 0.0001; Saliva: 269.0, 199.7−362.4 *vs* 53.1, 31.7−88.7; *p*< 0.0001) (Figure 2A). Subjects with a greater number of exposures to SARS-CoV-2 antigens showed higher antibody concentrations than those with a lower number of exposures (GMC BAU/mL, 95% CI, Blood: 6079, 4702−7859 *vs* 2033, 1384−2988; *p*< 0.0001; Saliva: 142.7, 84.7−240.4 *vs* 49.5, 28.9−84.5; *p*< 0.0001) (Figure 2B). Antibody concentrations decreased as the interval time between last exposure and sample collection increased (GMC BAU/mL, 95% CI, Blood: 6574, 4840−8929 *vs* 2326, 1673−3234; *p*< 0.0001; Saliva: 198.9, 129.6−305.2 *vs* 34.9, 19.9−61.2; *p*< 0.0001) (Figure 2C).

**Figure 2.**
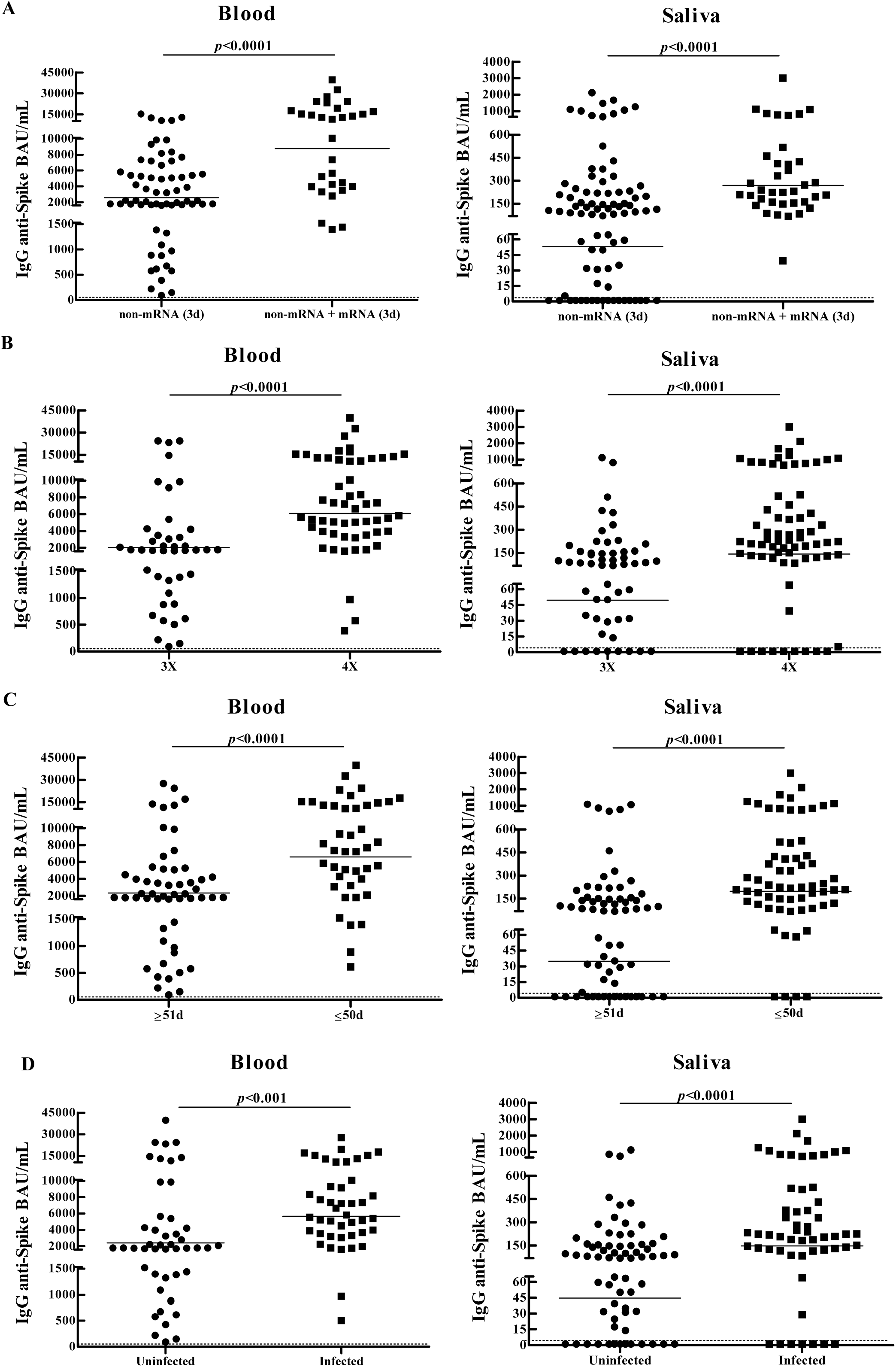
Comparison of systemic and salivary IgG antibody responses against SARS-CoV-2 in vaccinated adults. Blood and salivary anti-SARS-CoV-2 antibody levels were analyzed in vaccinated adults (*n* = 131) according to: vaccination schedules (A), number of exposures (B), time between last exposure and sample collection (C), SARS-CoV-2 infection (D). IgG anti-spike antibody concentrations (BAU/mL) with geometric means are shown. Dotted line indicates the assay detection limit (4.03 BAU/mL). *p* values were determined by Mann-Whitney test. Samples were assayed in duplicate and the results shown are representative of one of two independent experiments performed. non-mRNA (3d), three doses of non-mRNA-based-vaccines; non-mRNA + mRNA (3d), three doses combining mRNA and non-mRNA-based vaccines.

Adults who had been infected with SARS-CoV-2 showed higher antibody levels than uninfected subjects (GMC BAU/mL, 95% CI, Blood: 5660, 4379−7317 *vs* 2410, 1636−3549; *p*< 0.001; Saliva: 147.8, 84.1−259.8 *vs* 44.7, 27.0−73.9; *p*< 0.0001) (Figure 2D).

Using Fisher’s exact test, detection of specific antibodies in saliva was associated with schedules including mRNA-based vaccines (*p*< 0.001; OR=21.5), the time between last exposure and sample collection (*p*< 0.01; OR=4.7) and systemic antibody concentrations (*p*< 0.0001; OR=43.6).

### Comparison of IgG specific salivary antibody responses between vaccinated children and adults

Considering all samples, children showed higher salivary antibody concentrations compared to adults (GMC: 157.1 BAU/mL, 95% CI: 91.9−268.3 *vs* 79.8 BAU/mL, 95% CI: 54.9−115.9; *p*< 0.01). Therefore, antibody levels were compared between adults and children who shared the same conditions (vaccination schedules, number of exposures, time from last exposure, COVID-19 history). Children who received three doses of mRNA vaccines (mRNA (3d)) (GMC: 769.3 BAU/mL; 95% CI: 466.6–1268) or three doses combining mRNA and non-mRNA-based vaccines (non-mRNA + mRNA (3d)) (GMC: 1987 BAU/mL; 95% CI: 878.7–4492) showed higher antibody concentrations than adults who received three doses of schedules including mRNA-based vaccines (*p*< 0.001) (Figure 3A). Salivary antibody titers were higher in children compared to adults who had the same number of exposures (GMC BAU/mL, 95% CI, 3X: 195.0, 96.6−393.7 *vs* 49.5, 28.9−84.5; *p*< 0.0001; 4X: 744.6, 340.8–1627 *vs* 142.7, 84.7–240.4; *p*< 0.0001) (Figure 3B). Antibody concentrations were higher in children compared to adults who had the same interval time (≤58 days) between last exposure and saliva collection (GMC: 515.2 BAU/mL, 95% CI: 267.1−993.9 *vs* GMC: 174.0 BAU/mL, 95% CI: 113.1−267.8; *p*< 0.001) (Figure 3C). Children who had been infected with SARS-CoV-2 showed higher antibody levels than adults who had been infected (GMC BAU/mL, 95% CI: 783.2, 368.1−1667 *vs* 147.8, 84.1−259.8) (*p*< 0.01) (Figure 3D).

**Figure 3.**
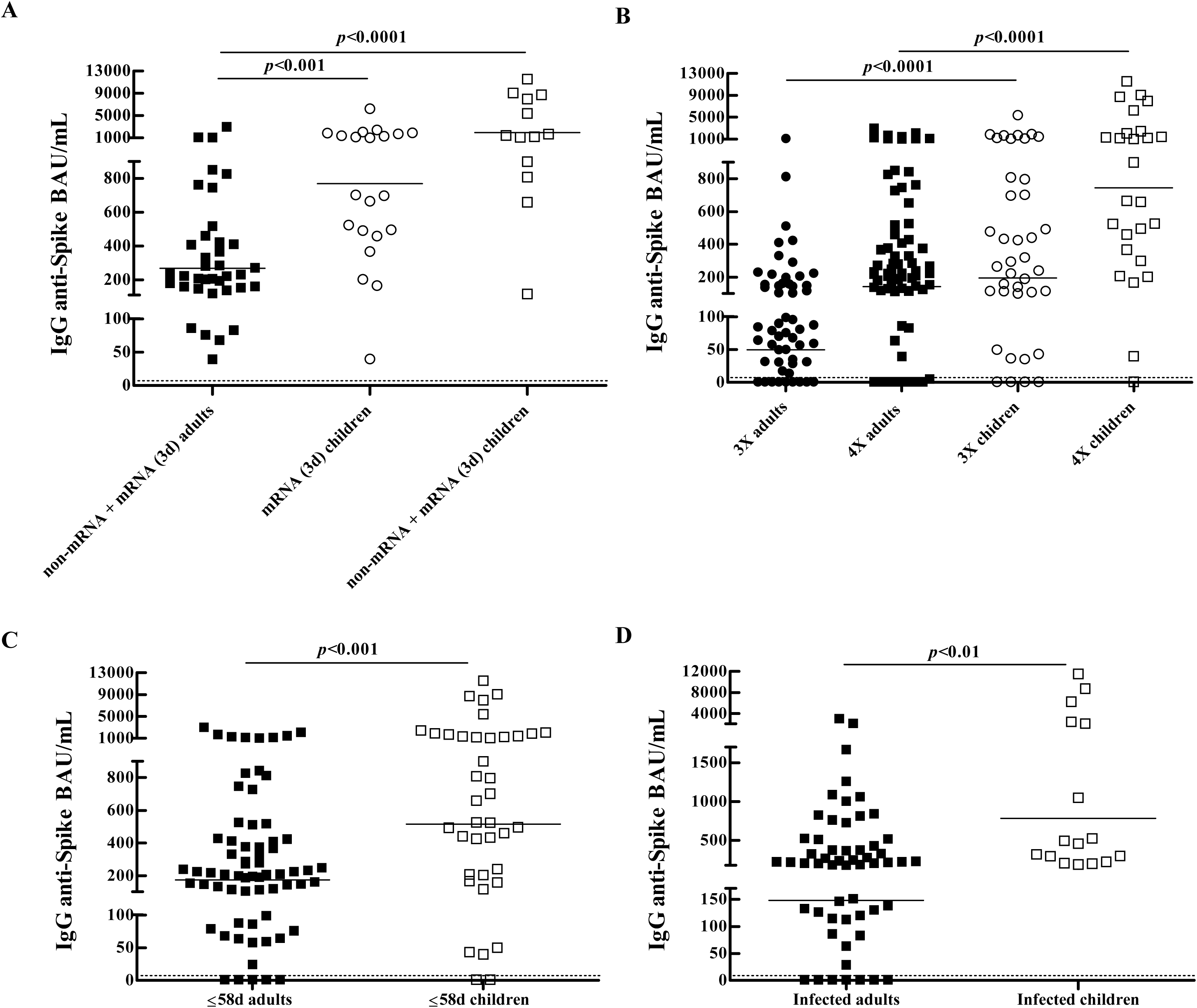
Comparison of salivary antibody responses against SARS-CoV-2 between vaccinated children and adults. Salivary anti-SARS-CoV-2 antibody levels were compared between vaccinated adults (*n* = 131) and children (*n* = 88) according to: vaccination schedules (A), number of exposures (B), time between last exposure and sample collection (C), SARS-CoV-2 infection (D). IgG anti-spike antibody concentrations (BAU/mL) with geometric means are shown. Dotted line indicates the assay detection limit (4.03 BAU/mL). *p* values were determined by Mann-Whitney test. Samples were assayed in duplicate and the results shown are representative of one of two independent experiments performed. non-mRNA + mRNA (3d), three doses combining mRNA and non-mRNA-based vaccines; mRNA (3d), three doses of mRNA-based vaccines.

Multivariable linear regression analysis showed that younger age (β1: 874.2, 95% CI: 468.6−1280; *p*< 0.0001), number of exposures (β2: 364.0, 95% CI: 3.6−724.3; *p*< 0.05), schedules including mRNA-based vaccines (β3: 560.6, 95% CI: 147.4−973.7; *p*< 0.01), and shorter time from last exposure (β4: 373.6, 95% CI: 3.5−748; *p*< 0.05) were associated with salivary antibody levels (p < 0.0001).

## Discussion

Data from studies comparing antibody responses against SARS-CoV-2 after infection or vaccination between children and adults are conflicting in blood [1–10] and in the mucosa [4, 15]. On the other hand, as studies comparing systemic and salivary humoral immune responses to SARS-CoV-2 have yielded contradictory results [11, 15, 16–22], it remains yet to be determined whether saliva could be used as a non-invasive alternative to blood for the determination of antibodies against SARS-CoV-2.

In this work, we investigated and compared the systemic and salivary IgG antibody responses against SARS-CoV-2 in vaccinated adults. Analysis of different variables previously reported to be related to systemic anti-SARS-CoV-2 IgG antibody levels showed similar results for blood and saliva. Higher antibody concentrations were observed in both the blood and saliva of vaccinated adults who had been infected with SARS-CoV-2, received schedules including mRNA-based vaccines, had more exposures, and a shorter interval time between last exposure and sample collection. Furthermore, a positive correlation was found between systemic and salivary anti-SARS-CoV-2 IgG antibody levels, with higher salivary antibody concentrations observed in subjects with higher systemic antibody levels. However, specific antibodies were detected in 100% of blood samples and in 84% of saliva samples. Detection of specific salivary antibodies was associated with schedules that included mRNA-based vaccines, time between last exposure and sample collection, and systemic antibody concentrations. These results, together with the fact that most subjects without detectable salivary antibodies showed lower systemic antibody levels, suggest that salivary antibody detection could depend on variables that influence systemic antibody levels. Therefore, although our results showed that the salivary immune response against SARS-CoV-2 in vaccinated adults largely reflects that observed at systemic levels, salivary antibody detection might not be possible in subjects presenting lower systemic antibody concentrations. This is in agreement with previous results showing that salivary antibody responses were mainly detected in subjects showing higher serum antibody levels [25]. However, in our studies, the prevalence of salivary antibodies was high in both adults (84%) and children (81%) [12], in contrast to previous results in unvaccinated infected subjects [11, 13, 25] but in agreement with studies in vaccinated subjects [13, 18, 22–23]. This suggests that multiple exposures are likely required to enhance systemic antibody levels and allow salivary antibody detection.

We also compared salivary antibody concentrations between vaccinated children and adults to analyze whether specific antibody responses differed within the salivary compartment. Vaccinated children showed a higher salivary antibody response against SARS-CoV-2 compared to adults. This difference remained when antibody levels were compared between vaccinated children and adults who shared the same conditions (vaccination schedules, number of exposures, time from last exposure, COVID-19 history). Multivariable linear regression analysis showed that younger age, number of exposures, schedules including mRNA-based vaccines, and shorter time from last exposure were associated with salivary anti-SARS-CoV-2 IgG antibody levels.

The biological and molecular basis for a favorable outcome after SARS-CoV-2 infection in children compared to adults is not fully understood. Several hypotheses have tried to explain the milder presentation of the disease in children, including a protective role of pre-existing cross-reactive antibodies against seasonal coronaviruses, a lower expression of the angiotensin-converting enzyme 2 (ACE2) and a less propensity to develop an exacerbated pro-inflammatory response, among others [26]. However, the contribution of the antibody response against SARS-CoV-2 to the favorable outcome in the pediatric population is not entirely clear. In the current work, we showed that children had higher salivary anti-SARS-CoV-2 IgG antibody concentrations than adults. Given that mucosal immunity plays a key role in prevention and early defense against infection, being the entry route for the virus and the site of first encounter with the immune response, it is tempting to speculate that the stronger salivary antibody response observed in children could help prevent or limit infection, thus contributing to the favorable outcome observed in the pediatric population.

Although salivary IgG is primarily derived from circulating IgG through transudation [27] and the salivary IgG response in adults highly reflects the systemic antibody response, it remains to be determined whether the salivary antibody response in children also similarly reflects that in blood. Besides, given there is also some local IgG production, it cannot be ruled out that a difference in the local antibody response mounted may also account to the observed difference in the salivary antibody response against SARS-CoV-2 between children and adults.

Limitations of this study included the lack of information on the systemic antibody response in children, which precluded the comparison between systemic and salivary immune responses in them. The limited availability of saliva samples (especially those from children that were diluted several times), prevented further investigation on neutralizing and specific IgA antibody responses. While some reports showed that salivary anti-SARS-CoV-2 IgG antibody levels strongly correlate with the neutralizing capacities of both serum [18] and saliva [23], knowledge of salivary IgA and neutralizing antibody responses could also add insights related to the local immune protection in children and adults. Information about SARS-CoV-2 variants involved in the infections/household contact exposures and their association with antibody levels was not available. However, most infections/household contact exposures, both in adults and children, occurred when the only circulating variant in Argentina was Omicron.

## Conclusions

Altogether, our results showed that the salivary IgG antibody response against SARS-CoV-2 largely reflects the systemic response in vaccinated adults. However, salivary antibodies may not be detectable in subjects with low systemic antibody levels. Nevertheless, since saliva sampling is non-invasive and allows self-collection, salivary antibody determination could be performed as an initial screening to evaluate the antibody response against SARS-CoV-2. Besides, as salivary antibody response is indicative of the systemic, salivary antibody determination could guide vaccination strategies by administering booster doses to subjects without detectable salivary antibodies, allowing them to achieve high systemic antibody levels.

In conclusion, the determination of salivary antibodies against SARS-CoV-2 could be a non-invasive approach for the evaluation of the short-term immune response in subjects with multiple exposures, either resulting from vaccination combined with infection or from the administration of immunogenic vaccination schedules, both in the adult and pediatric population, especially in settings where blood sampling cannot be performed.

Furthermore, the higher levels of salivary anti-SARS-CoV-2 IgG antibodies observed in children suggests that local immune protection may differ between children and adults. Whether the stronger salivary antibody response in children provides improved local immune protection and contributes to the favorable outcome in the pediatric population warrants further investigation.

## Data Availability

All data produced in the present study are available upon reasonable request to the authors

## Acknowledgments

This study was supported by a grant from CONICET (PIP 11220210100378CO). Some aspects of this work could not have been fulfilled without the generous contribution of the IIHEMA and the Academia Nacional de Medicina, who provide financial support to our ongoing research. The authors thank all the children and adults enrolled in this study for their participation and collaboration.

## Author contributions

M.N.B and P.B conceived and designed the study, M.N.B, I.K., M.P, N.A, F.S., and P.B.; participated in sample and clinical data collection, M.N.B, I.K., F.S., and P.B., performed the experiments, M.N.B and P.B.; analyzed and interpreted the data., M.N.B. and P.B.; drafted the first version and wrote the final version of the manuscript. All authors read and approved the final manuscript version.

## Abbreviations

BAU: binding antibody units
BAU/mL: per mL
GMC: geometric mean concentrations
95% CI: 95% confidence intervals.

